# The Effects of Coronavirus Victimization Distress and Coronavirus Racial Bias on Mental Health Among AIAN, Asian, Black, and Latinx Young Adults

**DOI:** 10.1101/2020.08.19.20178343

**Authors:** Celia B. Fisher, Xiangyu Tao, Tiffany Yip

**Affiliations:** Fordham University

## Abstract

**Rationale:** U.S. Racial/ethnic minorities have been disproportionately impacted by the COVID-19 pandemic in rates of infection and morbidity. Pre-pandemic racial discrimination has been associated with depression and general anxiety. However, the effect of Coronavirus specific forms of discrimination on mental health have not been examined. This study assessed the effect of previously identified social determinants of mental health and COVID-19 specific victimization and racial bias beliefs on depression and anxiety among young adults of color in the U.S.

**Methods:** A national online survey of 399 AIAN, Asian, Black, and Latinx adults (18 – 25 years) included demographic variables, COVID-19 health risks, and standardized measures of depression, anxiety, Coronavirus related victimization distress and perceptions of Coronavirus-related racial bias across a range of contexts.

**Results:** Employment, financial and prescription insecurity, COVID-19 health risks, Coronavirus victimization distress and Coronavirus racial bias beliefs were positively correlated with depression and anxiety. Scores on the Coronavirus racial bias scale were significantly higher among Asian and Black respondents. Structural equation modeling controlling for race/ethnicity and demographic variables indicated perceived Coronavirus racial bias mediated the effect of Coronavirus victimization distress on both mental health indices.

**Conclusion:** Results suggest the COVID-19 pandemic has created new pathways to mental health disparities among young adults of color by reversing formerly protective factors such as employment, and by exacerbating structural and societal inequities linked to race. Findings highlight the necessity of creating mental health services tailored to the specific needs of racial/ethnic minorities during the current and future health crises.

---

The COVID-19 pandemic has amplified existing racial/ethnic health disparities in the United States. American Indian/Alaskan Natives (AIAN), Asian, Black, and Latinx young and older adults are at disproportionally higher risk for COVID-19 infection and mortality (CDC, 2020a; Sze et al., 2020). These disparities are rooted in long-standing racial/ethnic inequities in medical and behavioral health treatment utilization, access to culturally relevant health services, treatment satisfaction and trust, and service outcomes (Ben et al., 2017; Chen, 2019; Manuel, 2018; Melillo, 2020). Although rates differ among racial/ethnic groups, compared with non-Hispanic Whites, AIAN, Asian, Black, and Latinx persons in the U.S. have higher rates of CDC-identified medical conditions associated with risk of severe illness from COVID-19, including obesity, diabetes, asthma, cardiovascular disease, and HIV (Bancks et al., 2017; Beavis et al., 2017; Cho et al., 2014; Flegal et al., 2016; Gold & Wright, 2005; Gordon et al., 2019). In addition to these longstanding disparities in healthcare, existing racial/ethnic disparities in financial insecurity, exacerbated by reallocation of pharmaceuticals for emergency care, has created barriers to prescriptions for health conditions related to COVID-19 risk, placing people in lower economic strata and racially segregated communities at additional higher risk (CDC, 2020b). Moreover, racially and ethnically marginalized adults in the U.S. are more likely to be employed in the health care work force or as frontline workers in industries such as food services, pharmacies, personal care and public transportation increasing their risk of COVID-19 exposure (Bureau of Labor Statistics, 2018; Gordon et al., 2019; Rho et al., 2020; Wang et al., 2020).

## Social Determinants of Mental Health

During the COVID-19 pandemic, depression and anxiety among AIAN, Asian, Black, and Latinx people have also increased (Baldwin et al., 2020; Chen, 2019; McKnight-Eily et al., 2021). Although pre-pandemic prevalence rates of major depression and generalized anxiety among these groups were often reported to be similar to or lower than non-Hispanic Whites, these findings have been attributed to the fact that people of color in the U.S. seek mental health clinical care at rates well below their need, face discrimination during diagnosis and treatment, suffer from misdiagnosis and clinician bias, and within their communities mental health may be shrouded by silence and shame (Baldwin et al., 2020; Harkness et al., 2020; Mental Heath America, 2021; Novacek et al., 2020; Vilsaint et al., 2019). Further, although prior research has repeatedly reported on the protective effect of employment on mental health (McGee & Thompson, 2015; Paul & Moser, 2009), during the pandemic, employed individuals, especially those whose work involves in-person contact, are experiencing higher levels of depression and anxiety (McKnight-Eily et al., 2021; Mehdi et al., 2020). Increases in externalizing disorders among racial/ethnic minority adults during the the pandemic, may also be a consequence of stress associated with pre-existing health conditions, work-related increases in exposure to COVID-19 infection, loss of a loved one to the virus, existing inequities in income and savings, and pandemic related fears of unemployment and food insecurity (Arenas et al., 2019; Donnelly & Farina, 2021; Mayo Clinic, 2020; McCurley et al., 2019; Nagata et al., 2019; Oh et al., 2019; Schachter et al., 2018).

## Racial/Ethnic Discrimination and Mental Health

AIAN, Asian, Black, and Latinx populations in the United States experience unique stressors related to their marginalized social identity that adversely contribute to depression and anxiety, including explicit racial/ethnic discrimination and microaggressions (Chin et al., 2020; Forrest-Bank & Cuellar, 2018; Lui, 2020). Intergenerational trauma created by enslavement of individuals of African descent and subsequent Jim Crow laws, colonialization of Indigenous people and violation of sacred lands, and labor-based exploitation and harsh immigration laws affecting Latinx and East Asian peoples has been shown to effect both physical and mental health among socially marginalized racial/ethnic groups (Barlow, 2018; Chu et al., 2020; Cobb et al., 2019; Farisi et al., 2019; Sandoiu, 2020). There is growing evidence that this has increased during the current pandemic (Addo, 2020; Dhanani & Franz, 2021; Ruiz et al., 2020).

Although to date, the effect of Coronavirus specific victimization and racial bias on mental health among U.S. racial/ethnic social minorities has not been examined, research on victimization distress in response to contagious diseases such as HIV, H1N1, and the 2003 severe acute respiratory syndrome (SARS) has found that both nationally and globally contagious disease discrimination is associated with long-term negative mental health outcomes including depression and anxiety (Crockett et al., 2019; Goodwin et al., 2009; Siu, 2008; Williams & Gonzalez-Medina, 2011). For young U.S. racial/ethnic social minorities living through the COVID-19 pandemic, the extent to which personal experiences with Coronavirus-related victimization are directly related to poor mental health outcomes may be influenced by perceptions regarding Coronavirus related racial bias across the country, and consequently, the extent to which they attribute Coronavirus victimization as racially based (Mouzon et al., 2017; Potter et al., 2019).

## The current study

Although the devastating impact of COVID-19 on the physical health of people of color in the U.S. has been well established, there is a paucity of research on how pre-existing health and current economic and social factors are affecting the mental health of these populations (Galanis & Hanieh, 2021). Public association of members of racial/ethnic groups in the U.S. as essential workers who are more likely to be infected with the Coronavirus, upsurges in racially based hate crimes and the COVID-19 specific tide of politically fomented related hate and violence against Asian Americans (McCarthy, 2020; Weiss, 2021) underscores the urgency of studying the effects of Coronavirus specific forms of discrimination and societal racial bias on the mental health of AIAN, Asian, Black, and Latinx young adults in the United States.

The primary aim of the current study was to examine the relationship between Coronavirus victimization distress and attributions of Coronavirus racial bias and the mental health of AIAN, Asian, Black and Latinx young adults during the first phase of the COVID-19 pandemic. The following hypotheses were tested: (1) Essential worker status, lower household income, financial and prescription insecurity, and number of COVID-19 health risks will be associated with higher levels of depression and anxiety; (2) Higher levels of Coronavirus victimization distress and perceptions of Coronavirus racial bias across healthcare, social media and other settings will be associated with higher levels of depression and anxiety; (3) Perceptions of Coronavirus racial bias will mediate the association between Coronavirus victimization distress and depression and anxiety among young adults of color when other variables are held constant.

## Methods

### Participants

Data were collected during April 2020 as part of a larger Internet-based national survey on the biological, psychological and social impact of the Coronavirus pandemic among young adults from diverse racial/ethnic groups. Eligible participants self-identified as AIAN, Asian, Black, or Latinx; were between 18 – 25 years of age; self-reported they did not have/had the Coronavirus; lived in the United States for more than one year; and could read English at an eighth-grade level.

### Procedure and data validation

Recruitment was conducted by Qualtrics XM which sent emails or posts to young adults who had signed up to take surveys across various survey panel websites and offered compensation worth $16.50 converted into the survey panels’ point systems and received within 7 days of completion. The emails and posts briefly described the study and provided a link to a screener on a different Qualtrics site. The screener included questions on participants’ age, race/ethnicity, assigned sex at birth, gender, sexual orientation, the U.S. state and zip code in which they lived, length of time living in the U.S., geographic region, living situation, employment and student status, and English reading proficiency.

The target goal was to recruit 100 participants from each of the four racial/ethnic groups, over-sampling individuals living in rural areas. A Qualtrics system feature excluded participants who did not meet eligibility criteria and prevented them from re-entering the screener. Manual data validation protocols were established to exclude fraudulent or repeat participants (e.g., consistency between age and date of birth; inconsistency between reported city in which the survey was taken and zip code). A speed check also excluded participants who responded in less than half the time of the median survey response. Approximately 57,000 people were contacted and a total of 223 AIAN, 198 Asian, 418 Black, and 195 Latinx participants completed the screener, with 535 meeting eligibility requirements. Of those 450 (84.11%) completed the survey and passed the speed check. An additional 51 respondents were eliminated because they reported having/had the Coronavirus and 7 were eliminated for missing data on the Coronavirus Racial Bias Scale, resulting in a final sample of 399. Primary racial/ethnic identity was reported as follows: AIAN (N = 86, representing 20 tribal memberships); Asian (N = 94; 54% East Asian, 46% South East Asian); Black (N = 128; 79% African American, 21% Caribbean, African, or Other); Latinx (N = 91; 47% Mexican, 18% Puerto Rican, 25% Central American, South American, or Other).

Following the screener, eligible individuals were sent to an informed consent page. Participants who selected “I agree” at the bottom of the page were redirected to the survey which consisted of 204 items (average completion time 14.02 minutes; SD = 33.84). Participants were able to quit the survey at any time by closing the survey window, these data were not included in analysis. At the end of the survey, online resources on health information and Coronavirus prevention were provided. Qualtrics uses unique identification numbers for each participant so that identifiable information and survey data are not stored; therefore, participants identity and contact information were unknown to the investigators. The study was approved by the University Institutional Review Board.

## Measures

### Demographic information

Demographic information collected included age, sex assigned at birth, gender, race and ethnicity, sexual orientation, employment (full or part-time; “essential worker” status), student status and education, region, household income, financial and prescription insecurity, and CDC COVID-19 health risks (see Tables 1 and 2).

**Table 1.**
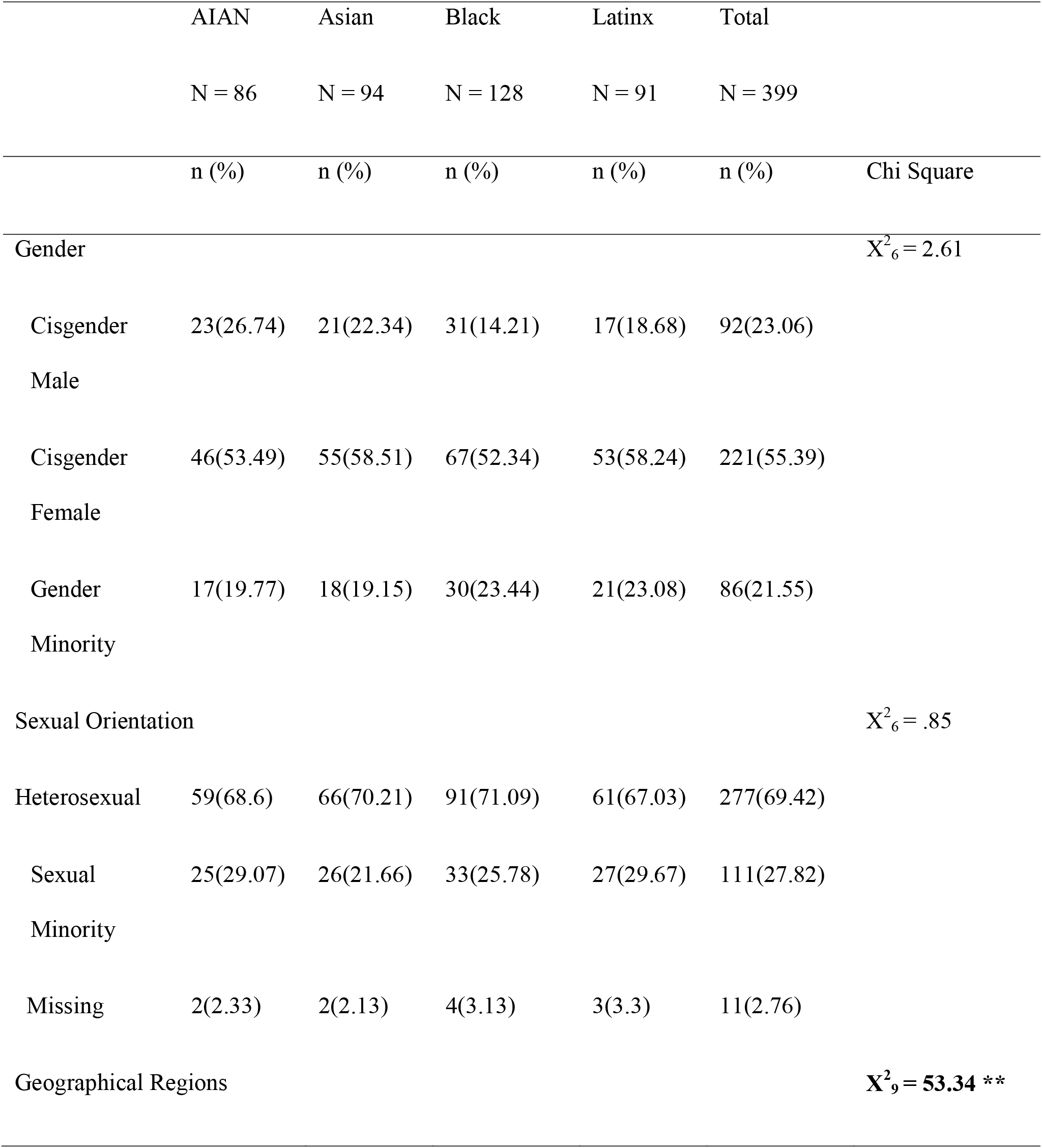
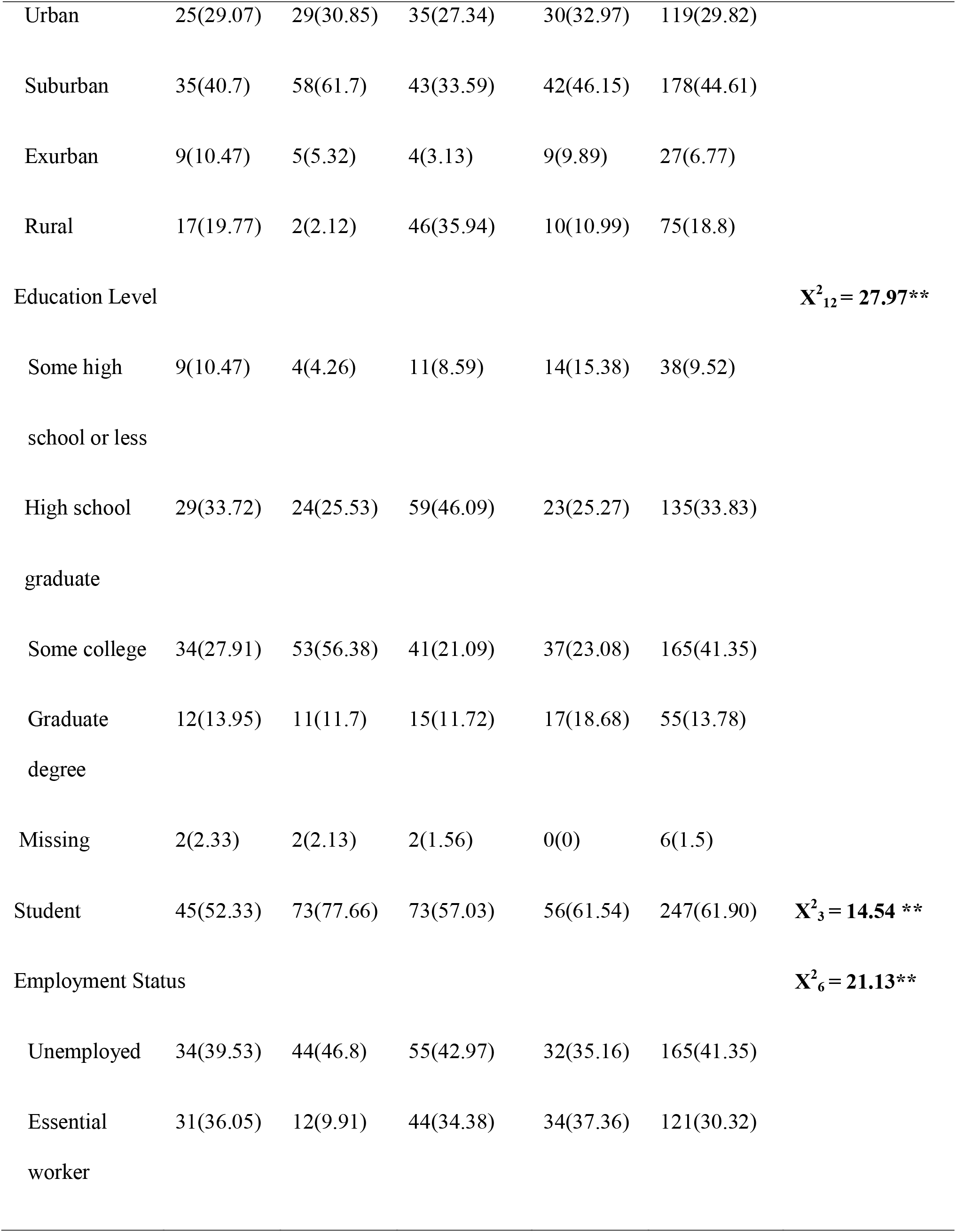
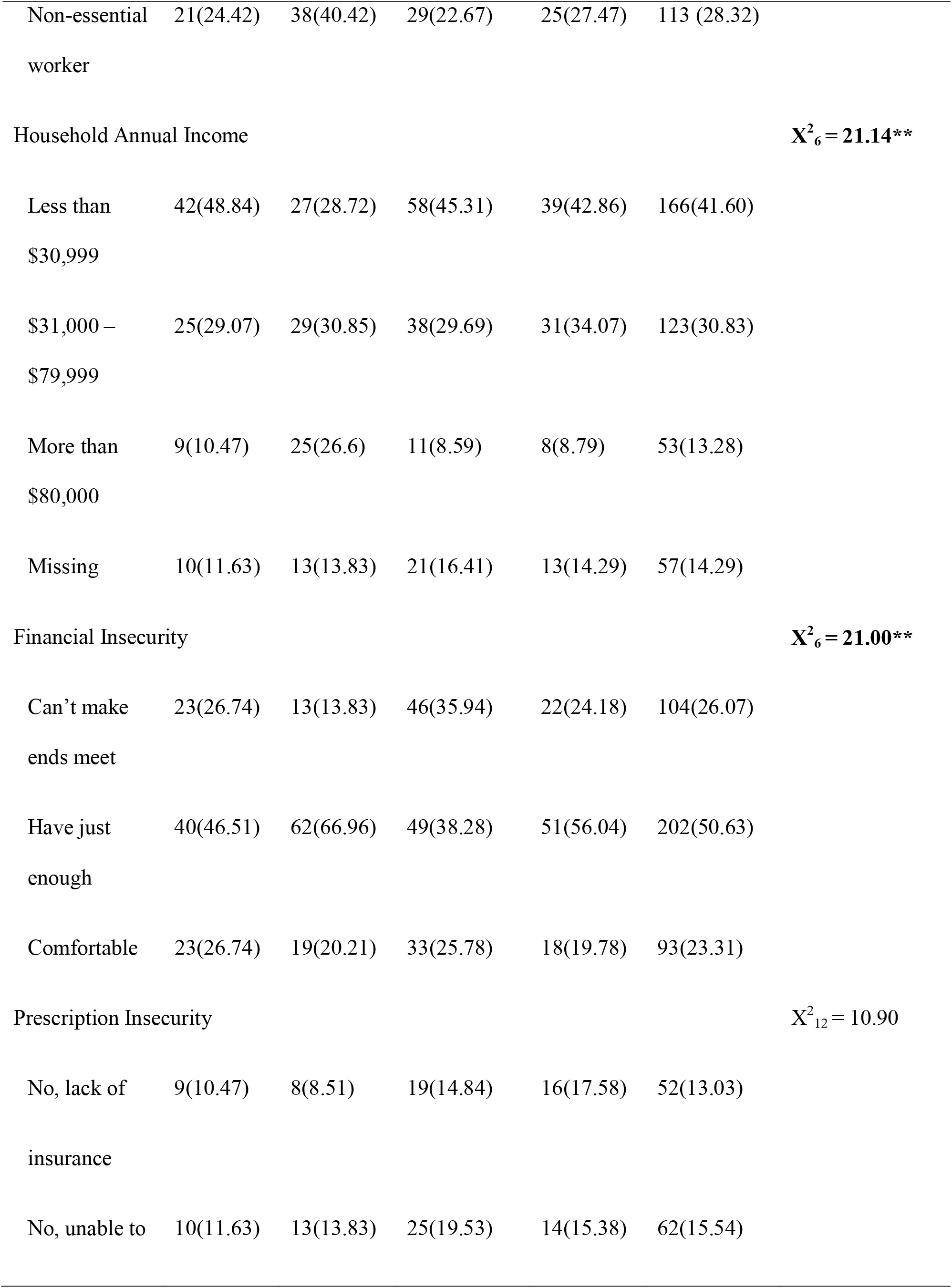
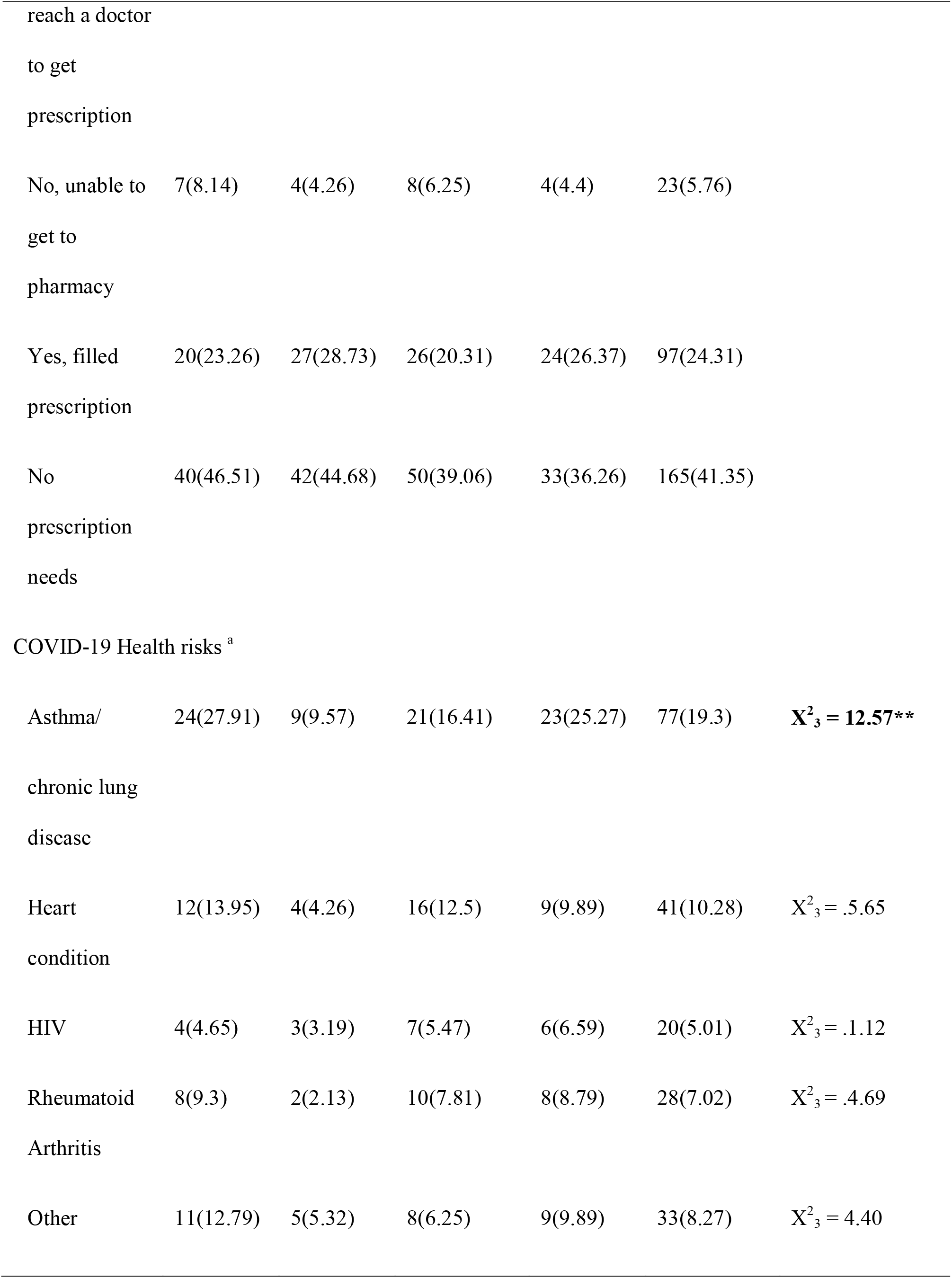
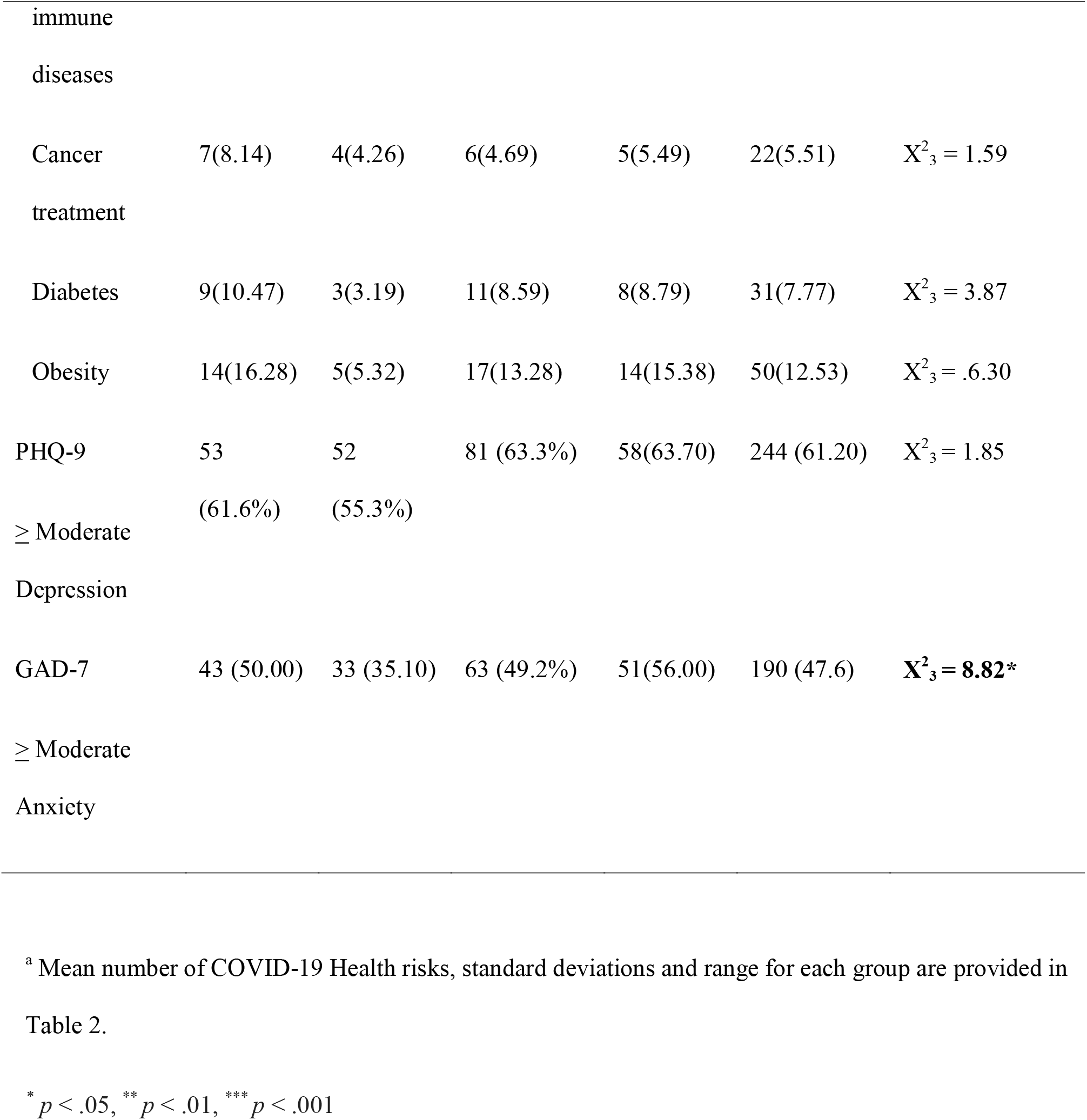
Frequency, Percentages and Chi Square Tests on Group Differences for Sociodemographic Characteristics, COVID-19 Health Risks, and Mental Health Screening Criteria

**Table 2.**
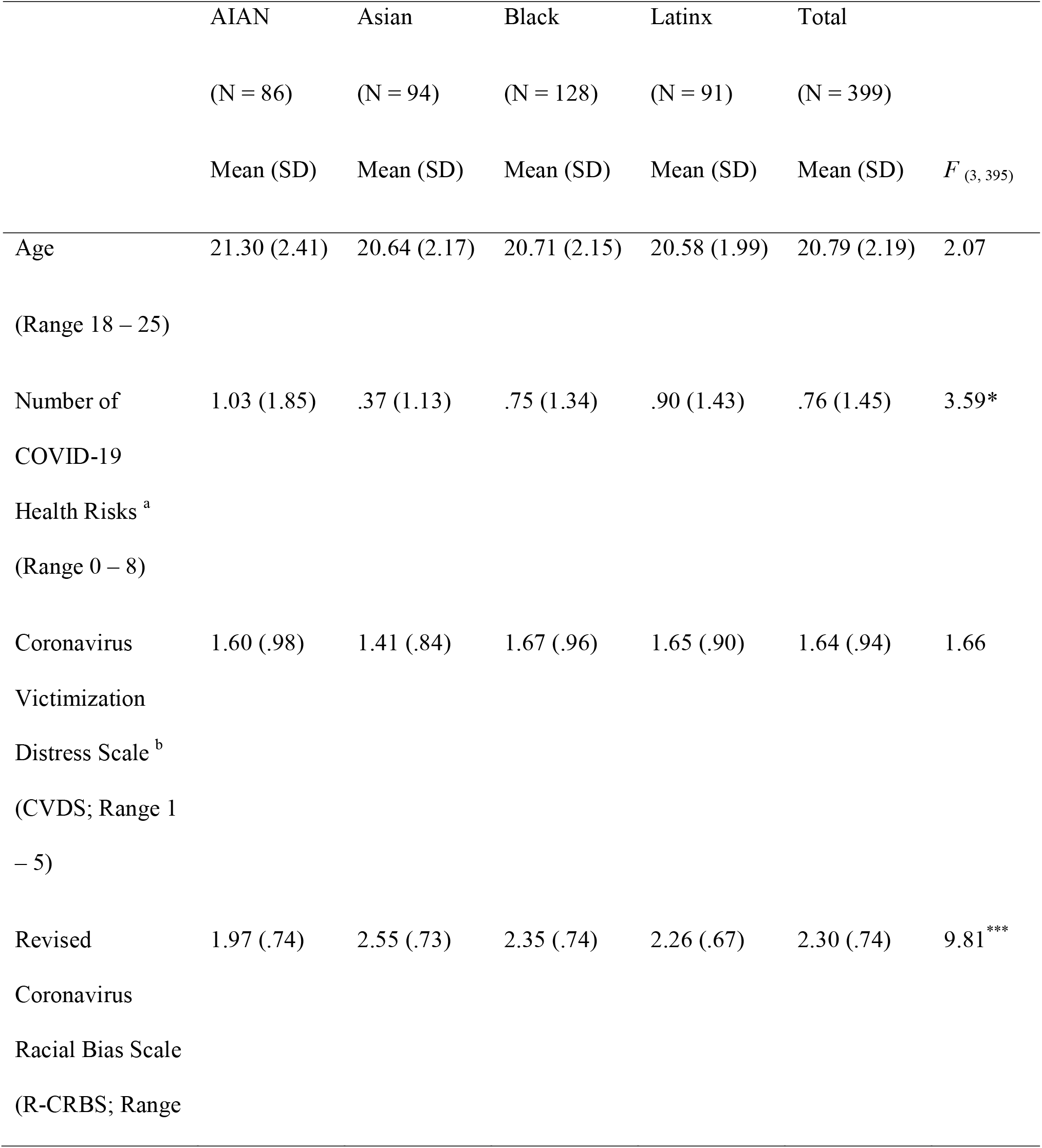
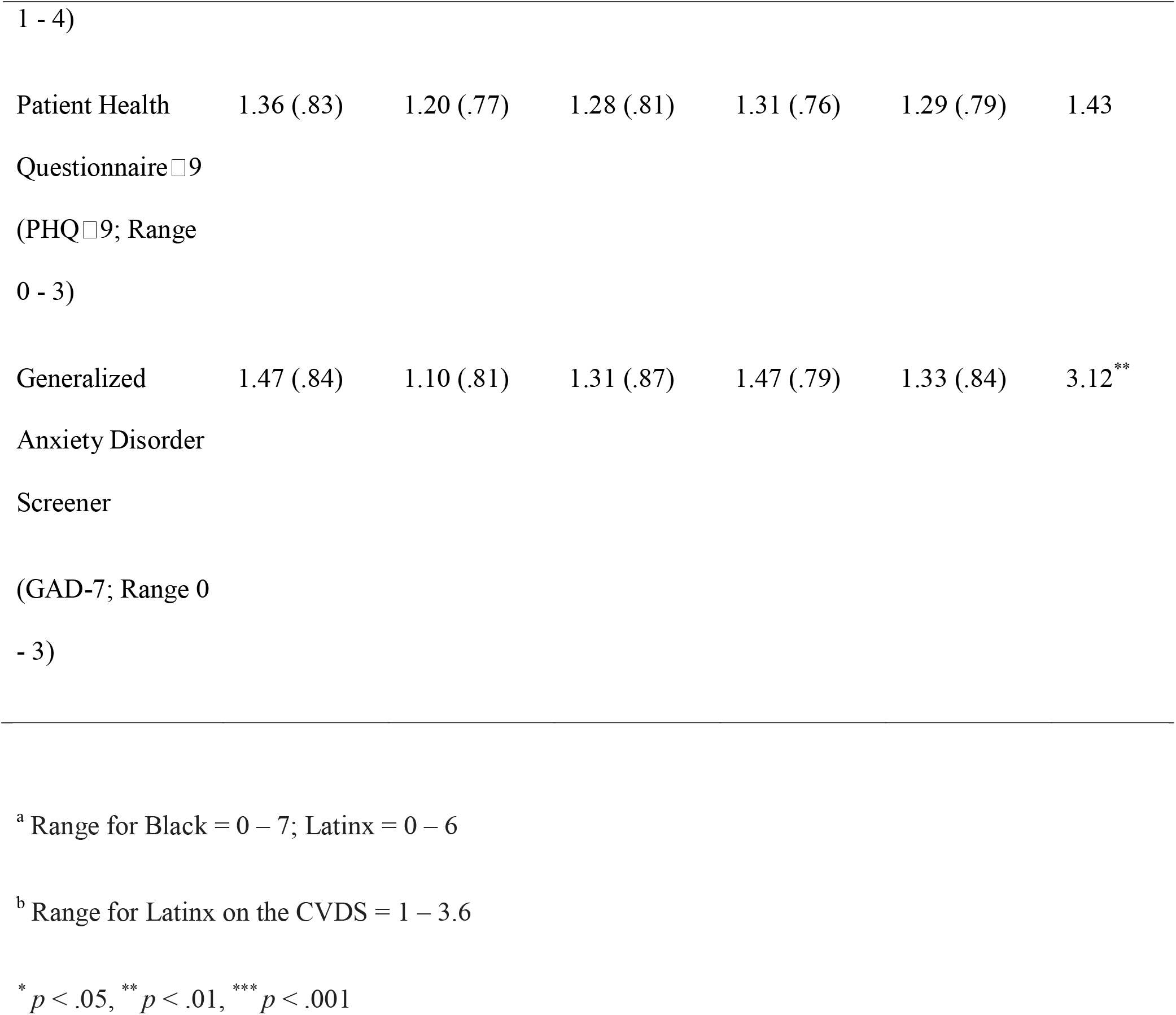
Means, Standard Deviations, Ranges and Racial/Ethnic Group Comparisons for Age, COVID-19 Health Risks, Coronavirus Victimization Distress, Coronavirus Racial Bias, Depression, and Anxiety

|  | AIAN | Asian | Black | Latinx | Total |  |
| --- | --- | --- | --- | --- | --- | --- |
|  | (N = 86) | (N = 94) | (N = 128) | (N = 91) | (N = 399) |  |
| | Mean (SD) | Mean (SD) | Mean (SD) | Mean (SD) | Mean (SD) | $F_{(3, 395)}$ |
| Age | 21.30 (2.41) | 20.64 (2.17) | 20.71 (2.15) | 20.58 (1.99) | 20.79 (2.19) | 2.07 |
| (Range 18 – 25) |  |  |  |  |  |  |
| Number of COVID-19 Health Risks <sup>a</sup> | 1.03 (1.85) | .37 (1.13) | .75 (1.34) | .90 (1.43) | .76 (1.45) | 3.59* |
| (Range 0 – 8) |  |  |  |  |  |  |
| Coronavirus Victimization Distress Scale <sup>b</sup> | 1.60 (.98) | 1.41 (.84) | 1.67 (.96) | 1.65 (.90) | 1.64 (.94) | 1.66 |
| (CVDS; Range 1 – 5) |  |  |  |  |  |  |
| Revised Coronavirus Racial Bias Scale | 1.97 (.74) | 2.55 (.73) | 2.35 (.74) | 2.26 (.67) | 2.30 (.74) | 9.81*** |
| (R-CRBS; Range |  |  |  |  |  |  |
| Patient Health | 1.36 (.83) | 1.20 (.77) | 1.28 (.81) | 1.31 (.76) | 1.29 (.79) | 1.43 |
| Generalized | 1.47 (.84) | 1.10 (.81) | 1.31 (.87) | 1.47 (.79) | 1.33 (.84) | 3.12<sup>**</sup> |

<sup>a</sup> Range for Black = 0 – 7; Latinx = 0 – 6
<sup>b</sup> Range for Latinx on the CVDS = 1 – 3.6
\* $p < .05$ , \*\* $p < .01$ , \*\*\* $p < .001$

### Coronavirus Victimization Distress Scale (CVDS)

The Coronavirus Victimization Distress Scale is a 5-item scale adapted from the Daily Heterosexist Experiences Questionnaire (Balsam et al., 2013), LGBT People of Color Microaggressions Scale (Balsam et al., 2011), and Adolescent Discrimination Distress Index (Fisher et al., 2000). The items assess distress in response to different instances of victimization, e.g., “because someone thought I was infected with the Coronavirus” (see Table 3). Responses ranged from 1 = It never happened, 2 = It happened but did not upset me; 3 = It happened and upset me a little; 4 = It happened and upset me moderately; 5 = It happened and upset me quite a bit. Cronbach’s alphas for this sample were computed following psychometric validation (see Data Analysis Plan and Results Section).

**Table 3.**
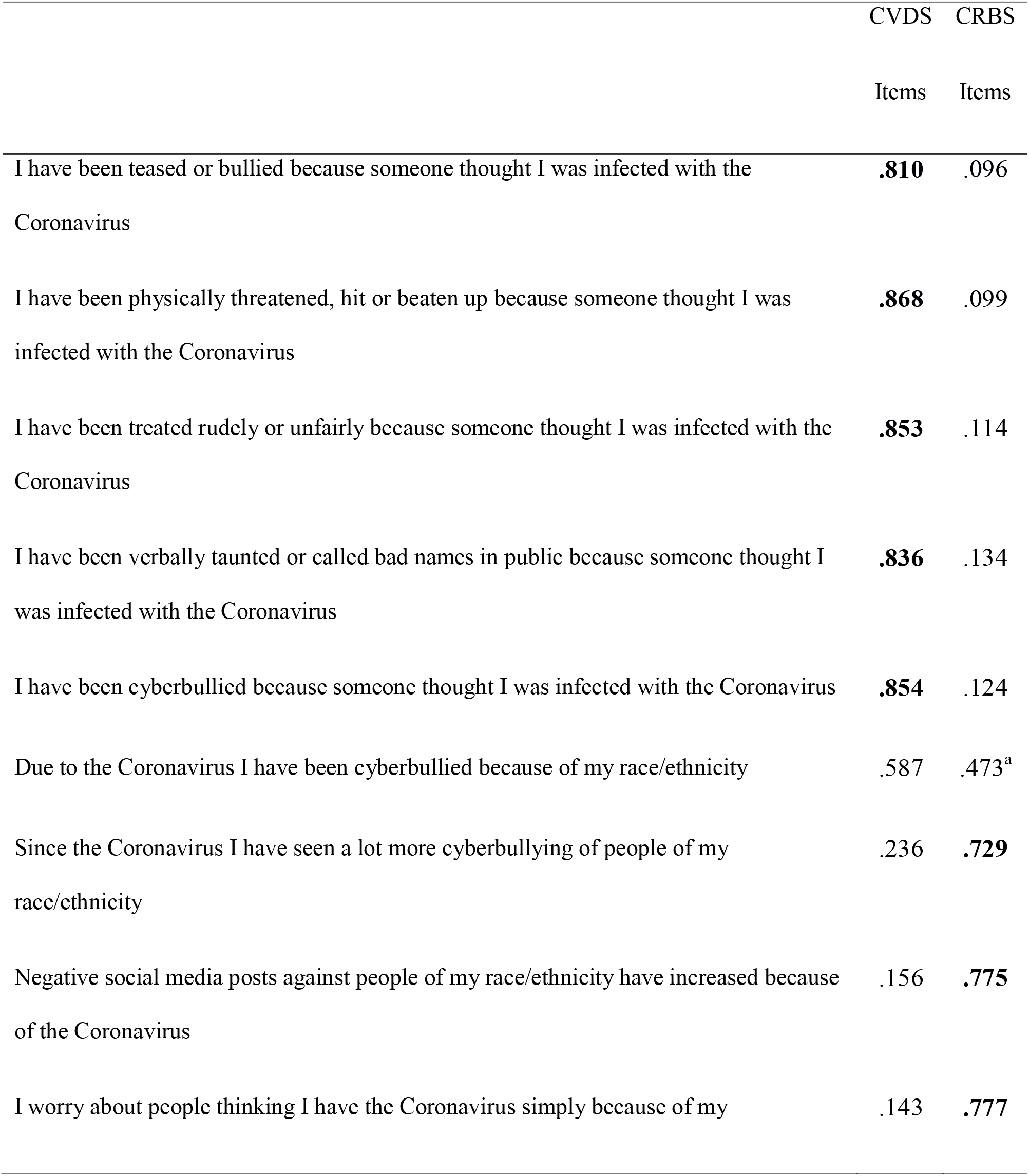
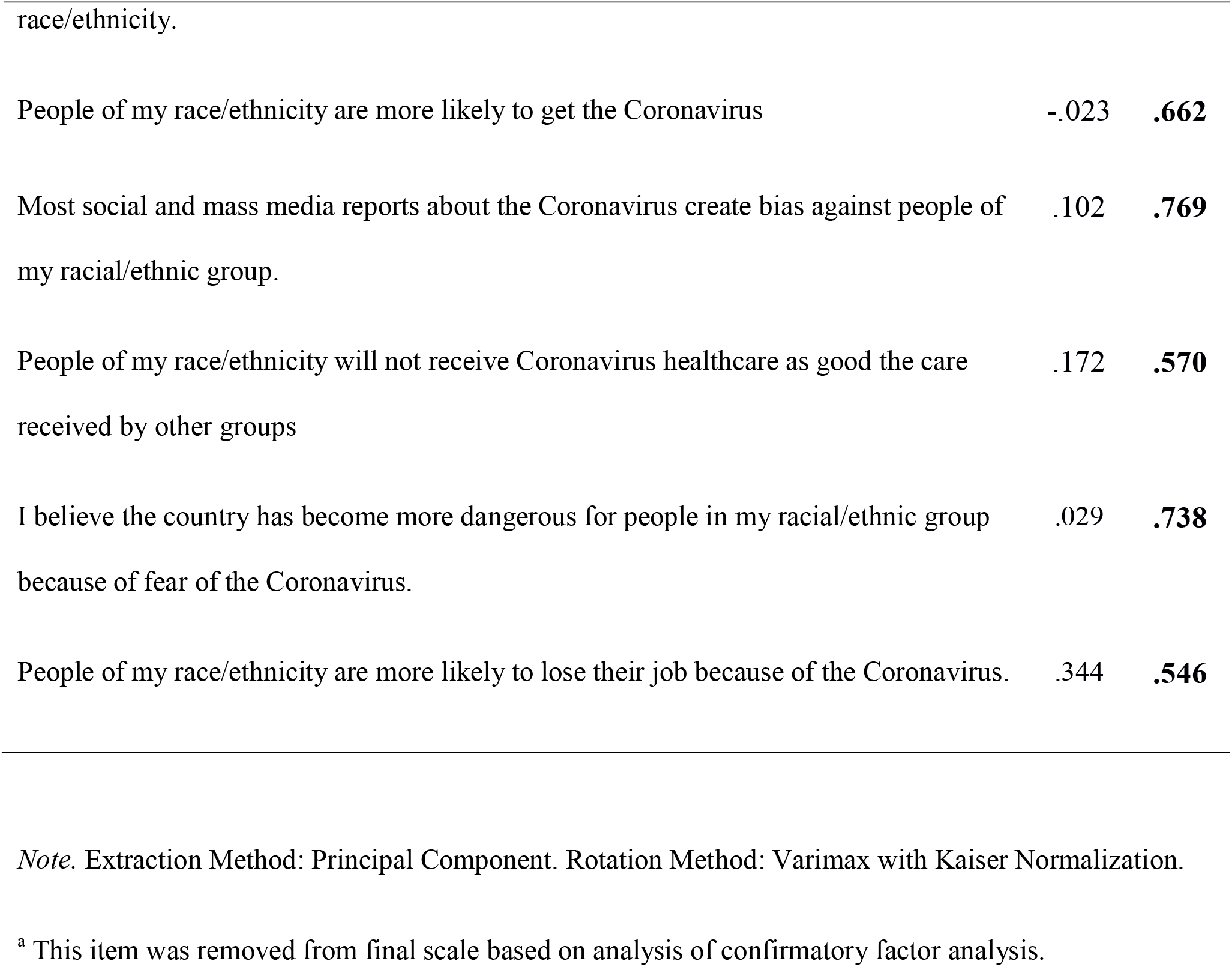
Factor Loadings for Coronavirus Victimization Distress Scale (CVDS) and Coronavirus Racial Bias Scale (CRBS) Based on CFA Model 1.

|  | CVDS | CRBS |
| --- | --- | --- |
|  | Items | Items |
| I have been teased or bullied because someone thought I was infected with the Coronavirus | **.810** | .096 |
| I have been physically threatened, hit or beaten up because someone thought I was infected with the Coronavirus | **.868** | .099 |
| I have been treated rudely or unfairly because someone thought I was infected with the Coronavirus | **.853** | .114 |
| I have been verbally taunted or called bad names in public because someone thought I was infected with the Coronavirus | **.836** | .134 |
| I have been cyberbullied because someone thought I was infected with the Coronavirus | **.854** | .124 |
| Due to the Coronavirus I have been cyberbullied because of my race/ethnicity | .587 | .473<sup>a</sup> |
| Since the Coronavirus I have seen a lot more cyberbullying of people of my race/ethnicity | .236 | **.729** |
| Negative social media posts against people of my race/ethnicity have increased because of the Coronavirus | .156 | **.775** |
| I worry about people thinking I have the Coronavirus simply because of my | .143 | **.777** |
| People of my race/ethnicity are more likely to get the Coronavirus | -.023 | **.662** |
| Most social and mass media reports about the Coronavirus create bias against people of my racial/ethnic group. | .102 | **.769** |
| People of my race/ethnicity will not receive Coronavirus healthcare as good the care received by other groups | .172 | **.570** |
| I believe the country has become more dangerous for people in my racial/ethnic group because of fear of the Coronavirus. | .029 | **.738** |
| People of my race/ethnicity are more likely to lose their job because of the Coronavirus. | .344 | **.546** |

*Note.* Extraction Method: Principal Component. Rotation Method: Varimax with Kaiser Normalization.
<sup>a</sup> This item was removed from final scale based on analysis of confirmatory factor analysis.

### Coronavirus Racial Bias Scale (CRBS)

The Coronavirus Racial Bias Scale is a 9-item scale developed to assess participants’ beliefs on whether the pandemic is negatively affecting societal attitudes toward one’s race/ethnicity, e.g. “I believe the country has become more dangerous for people in my racial/ethnic group because of fear of the Coronavirus” (see Table 3). Participants responded on a 4-point Likert-type scale (1 = Strongly disagree, 4 = Strongly agree). Cronbach’s alphas for this sample were computed following psychometric validation (see Data Analysis Plan and Results Section).

### Patient Health Questionnaire (PHQ-9)

The 9-item Patient Health Questionnaire (PHQ-9) assesses the frequency of past-month experiences with depressive symptoms (Kroenke et al., 2001). Sample items include “Trouble falling asleep, staying asleep, or sleeping too much” and “Feeling down, depressed or hopeless.” Items are scored on a 4-point scale: 0 = Not at all – 3 = Nearly every day. Prior Cronbach’s alpha for this scale ranged from .79 to .91 across AIAN, Asian, Black, and Latinx adults (Easton et al., 2019; Huang et al., 2006). For this sample, Cronbach’s alpha indicated good inter-item reliability (total sample = .89; range across racial/ethnic groups = .89 - .91).

### General Anxiety Disorder Screener (GAD-7)

The 7-item Generalized Anxiety Disorder Screener (GAD-7) (Spitzer et al., 2006) assesses anxiety symptoms during the past month, e.g. “Being so restless that it is hard to sit still” and “Feeling afraid as if something awful might happen.” Items are scored on a 4□point scale: 0 = Not at all, 3 = Nearly every day. In prior studies, Cronbach’s alpha for GAD-7 ranged from .79 to .91 across adults identifying as AIAN, Asian, Black, and Latinx (Dear et al., 2011; Löwe et al., 2008; Parkerson et al., 2015; Saunders, 2016; Wurster et al., 2019). For this sample, Cronbach’s alpha indicated good inter-item reliability (total sample =.90; range across racial/ethnic groups = .86 - .93.

### Data analysis plan

Descriptive statistics were calculated for demographic variables and mental health indices followed by analysis of variance (ANOVA) and Chi-square tests to assess racial/ethnic differences. To determine the independence and structure of the CVDS and CRBS, confirmatory factor analyses (CFA) were conducted followed by Cronbach alphas to assess resulting scale reliability. Correlational analyses and ANOVAs then examined associations among demographic variables, the CVDS, CRBS and mental health indices. Structural equation modeling (SEM) using the R-4.0.1. and *lavaan* package (R Core Team, 2020; Rosseel et al., 2017) assessed the hypothesis that Coronavirus racial bias mediates the effect of Coronavirus victimization distress on depression and anxiety. An alternative model testing whether Coronavirus victimization distress mediates the relationship between the mental health measures was also conducted. Goodness of fit indices included the comparative fit index (CFI), Tucker-Lewis Index (TLI), and the root-mean-square error of approximation (RMSEA). A fit of > .90 or .95 for the CFI and TLI and < .06 for RMSEA was considered adequate fit (Hu & Bentler, 1999). To test the indirect effects for statistical significance, the bias-corrected bootstrapping approach was adopted as it is robust against the violation of normal distribution assumptions for both the sampling distribution and indirect effect (MacKinnon et al., 2004); 1,000 re-samples were drawn to estimate the standard errors of the indirect effects and their 95% confidence intervals.

## Results

### Descriptive Data

Demographic data, percentages and Chi square tests of significance across each racial/ethnic group and total sample are provided in Table 1. The majority of the sample identified as cisgender females or males, heterosexual, were full or part-time employed (including 30.32% essential workers); and approximately half were students. The majority of respondents (N = 276; 69%) lived with their parents, a spouse or romantic/sexual partner. Almost half the sample reported an annual household income below or just above the poverty line ($30,680; (HHS, 2020). Slightly more than a quarter indicated they felt financially insecure (“Can’t make ends meet”) and 34% reported difficulty filling prescriptions during the past month. Asian respondents compared to the other racial/ethnic groups were less likely to be an essential worker and to report financial insecurity, were more likely to be a student, and reported higher household income and education; Black respondents were more likely than other groups to live in rural areas. Thirty-five percent reported at least 1 COVID-19 health risk (range = 0 – 8). Higher percentages of Black and Latinx young adults reported asthma or chronic lung disease. ANOVA followed by Tukey HSD tests indicated AIAN reported the highest and Asians reported the lowest number of health risks (see Table 2). There were no racial/ethnic age differences.

### Relationships among Demographic Variables and Mental Health Indices

Approximately 60% of respondents met screening criteria for moderate depression as assessed by the PHQ-9 (≥ = 10; (Kroenke et al., 2001); and close to 50% of respondents met GAD-7 screening criteria for moderate levels of anxiety (≥ = 10; Spitzer et al, 2006). Although there were no racial/ethnic differences for depression, Asian respondents reported significantly lower levels of anxiety than other groups (see Tables 1 & 2).

Correlations among variables are provided in Table 4. Consistent with hypothesis 1, financial and prescription insecurity and number of COVID-19 health risks were significantly associated with depression and anxiety measures. Both essential and non-essential workers had significantly higher scores on the mental health indices compared to unemployed respondents, but did not differ from one another (Tukey HSD tests following *F*_2, 396_ = 3.93, *p* < .001 and *F*_2, 396_ = 4.71, *p* < .001 for depression and anxiety, respectively). A combined employed/unemployed variable was significantly correlated with both mental health indices. Sexual minority participants reported higher levels of depression and anxiety than their heterosexual counterparts (*F*_1, 386_ = 28.60, *p* < .001 and *F*_1, 386_ = 17.68, *p* < .001, respectively) and gender minority respondents reported significant higher levels of depression than cisgender males, but not cisgender females (Tukey HSD test following *F*_2, 396_ = 3.68, *p* < .05). There was no effect of gender on anxiety nor any significant effects of age or household income on mental health indices.

**Table 4.**
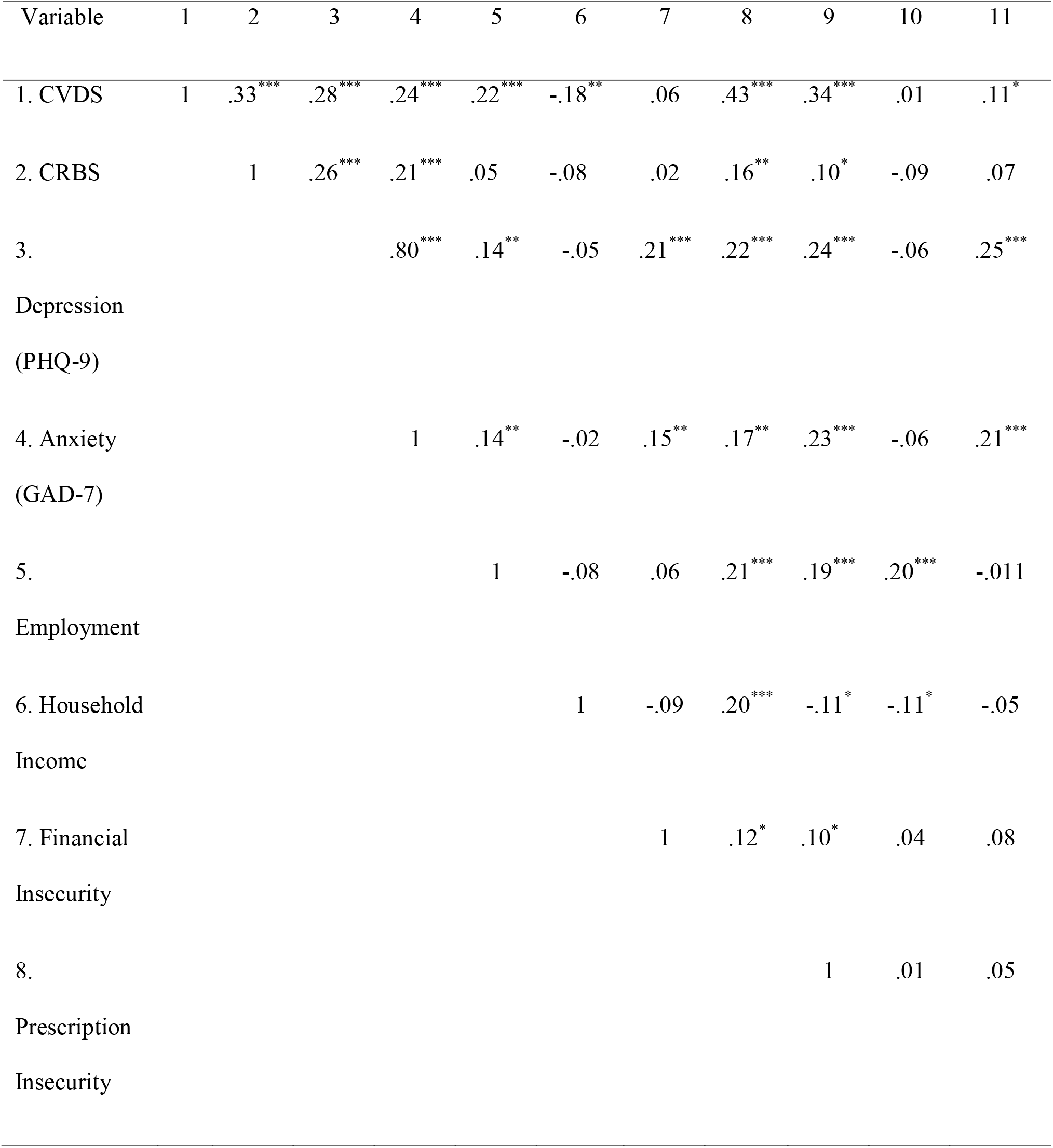
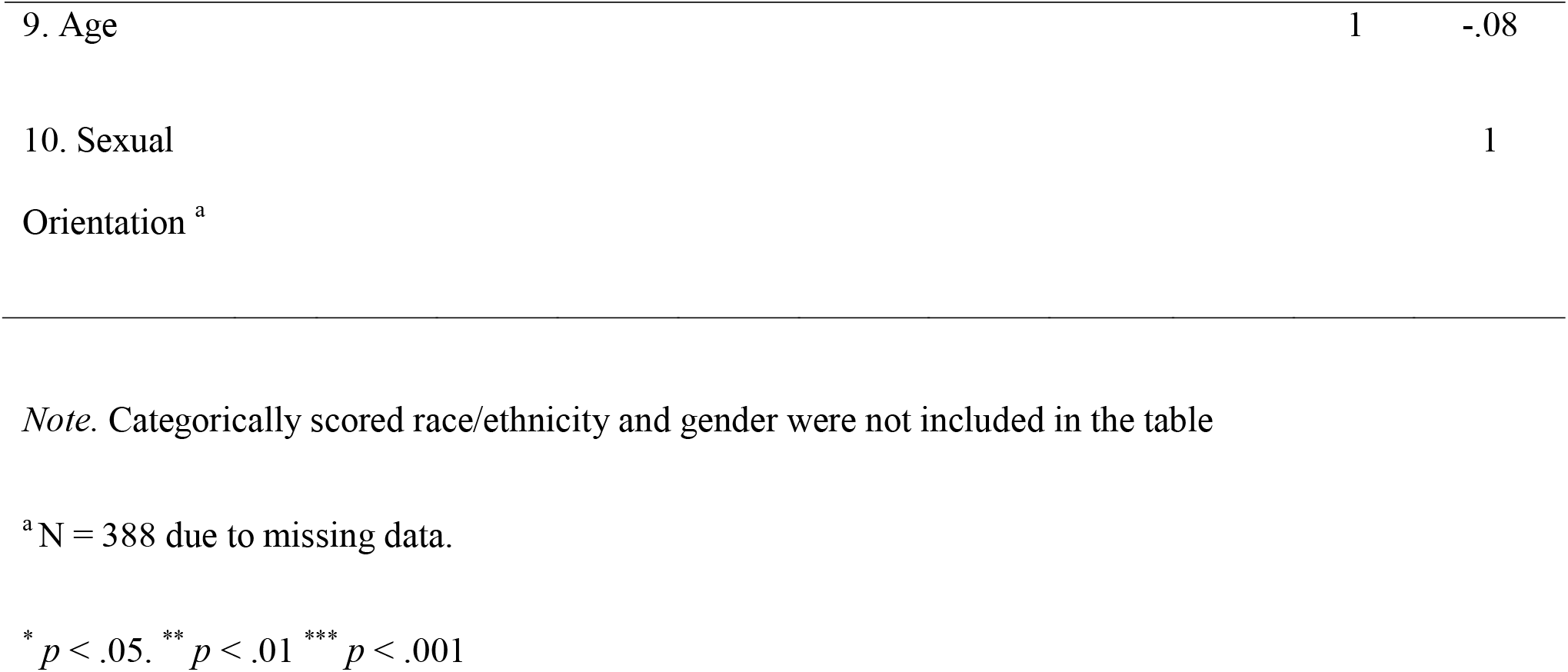
Correlation Matrix for Coronavirus Victimization Distress (CVDS), Coronavirus Racial Bias (CRBS), Depression, Anxiety and Covariates (N = 399).

### Psychometric Validation of the Coronavirus Victimization Distress Scale (CVDS) and Coronavirus Racial Bias Scale (CRBS)

Prior to conducting correlational and SEM analyses, three confirmatory factor analyses were conducted to determine the independence and factor structure of the CVDS and CRBS. Model 1 included all items on both scales (CFI = .90, TLI = .88, RMSEA = .097). Modification indices (MI = 90.65) indicated a strong correlated error between item 7 on the CRBS (“Due to the Coronavirus I have been cyberbullied because of my race/ethnicity”) and the CVDS (MacCallum, 1986); therefore this item was removed from further analyses (see Table 4). Model 2 yielded a relatively good fit between the 5-item CVDS and 8-item CRBS (CFI = .94, TLI = .92, RMSEA = .079). Cronbach’s alpha for the total sample for the CVDS was .91 (range = .90 - .93) and .87 (range = .84 - .87) for the 8-item CRBS. The two scales were positively correlated (*r* = .33, *p* < .001). To yield higher reliability and communalities for the SEM (Little et al., 2002), Model 3 was developed, with all 5 items on CVDS and four parcels constructed from the 8-item CRBS (created by randomly pairing two items together and calculating the mean). This model yielded the best fit to the data, CFI and TLI = .98, RMSEA = .06 (Hu & Bentler, 1999).

### Racial/Ethnic and Demographic Differences on the CVDS and CRBS

Mean scores and significant tests on the CVDS and CRBS among the 4 racial/ethnic groups are provided in Table 2. An ANOVA followed by Bonferroni post-hoc comparison tests (*p* = .001) indicated Asian and Black participants had significantly higher scores on the CRBS than AIAN and Latinx respondents, but no significant racial/ethnic differences emerged for the CVDS. Employed workers scored significantly higher on the CVDS than unemployed participants (*F*_1,397_= 19.24, *p* < .001); although employment was not significant for scores on the CRBS. Participants who had difficulty filling prescriptions compared to those reporting no prescription problems scored higher on both the CVDS (*F*_1,397_ = 87.47, *p* < .001) and CRBS (*F*_1,397_ = 9.94, *p* < .001). Gender minority persons reported higher scores on the CVDS (*F*_2,396_ = 7.26, *p* = .001; Tukeys HSD Test) than cisgender females and males. Pearson’s correlation tests indicated significant positive correlations between the number of COVID-19 health risks and scores on the CVDS and CRBS (Table 4). There were no differences on CVDS and CRBS based on age, sexual orientation, financial security, or annual household income.

### Tests of the Mediation Hypothesis

Consistent with hypothesis 2, scores on the CVDS and CRBS were significantly and positively associated with the depression and anxiety measures (Table 4). Structural equation modeling (SEM) was conducted to test the third hypothesis that perceived Coronavirus racial bias mediates the effect of Coronavirus victimization distress on depression and anxiety. Age, sexual orientation, financial security, COVID-19 health risks, prescription insecurity, and employment status (employed vs. unemployed) were included as covariates due to significant associations with the study variables. Based on the higher levels of anxiety reported by Asian respondents, racial/ethnic differences on anxiety were assessed using 3 dummy variables with Asian as the reference group. Gender differences emerged for depression, as such, 2 dummy variables were created with gender minority status as the reference group. Missing data (N = 11) on sexual orientation were handled by listwise deletion (Rosseel et al., 2017).

The primary analysis, which tested Coronavirus racial bias as mediating the effect of Coronavirus victimization distress on depression and anxiety is presented in Figure 1. The model yielded adequate fit on all three fit indices including the RMSEA (.052), 90% CI [.043 .061], CFI (.95), and TCL (.93) and found Coronavirus victimization distress had significant indirect effects on depression and anxiety (ß = .074, *p* =.003, 95% CI [.017, .081], ß = .074, *p* =.002, 95% CI [.019, .089], respectively); while direct effects non-significant. Consistent with hypothesis 3, the analysis indicated Coronavirus racial bias beliefs fully accounted for the influence of Coronavirus victimization distress on both mental health indices.

**Figure 1.**
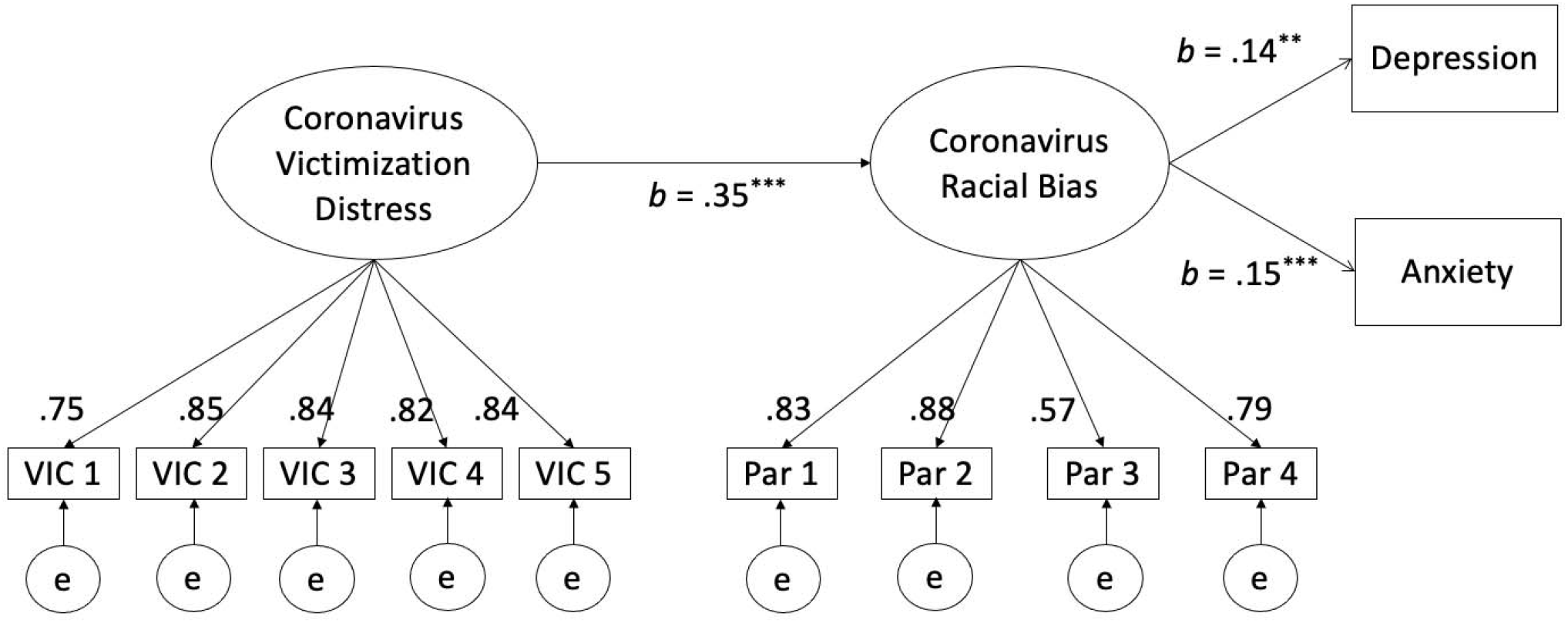
Standardized Results for Main Structural Equation Model with Bootstrapping Approach Testing the Mediating Effect of Coronavirus Racial Bias on the Association between Coronavirus Victimization Distress and Depression and Anxiety *Note*. Covariates included race/ethnicity, age, gender, sexual orientation, employment status, financial and prescription insecurity, and COVID-19 health risks. ^*^*p* < .05, ^**^*p* < .01, ^***^*p* < .001

### Test of Alternative Hypothesis

A third SEM analysis tested whether Coronavirus victimization distress mediated the effect of Coronavirus racial bias on depression and anxiety. We found an acceptable fit based on values of CFI, but the TLI (.88) and RMSEA (.07) did not support adequate fit nor were the standardized direct, indirect, and total effects significant.

## Discussion

AIAN, Asian, Black and Latinx people in the United States have been disproportionately impacted by the COVID-19 pandemic in rates of infection and morbidity. Explanations for these disparities include over-representation as essential workers and long-standing inequities in financial security and access to health services associated with racial/ethnic discrimination. Prior to the pandemic, racial/ethnic discrimination has been associated with poor mental health. Our data demonstrate that in addition to employment and health risks, victimization experiences and perceived increases in systemic racial biases specifically associated with the COVID-19 pandemic are related to higher levels of depression and anxiety among racial/ethnic minority young adults across geographic regions of the U.S.

Our results are consistent with recent research indicating higher levels of pre-existing health risks and financial insecurity among racial/ethnic minority groups are associated with increased internalizing disorders during the pandemic (Baldwin et al., 2020; Chen, 2019; Longmire-Avital, 2018; McKnight-Eily et al., 2021), and also demonstrates that prescription insecurity is contributing to mental health risk beginning at the first large scale wave of the virus in 2020. Prior studies have found employment to be a protective factor against mental health problems (McGee & Thompson, 2015; Paul & Moser, 2009). The present study also extends to racial/ethnic minority populations recent reports that COVID-19 pandemic represents a unique situation in which employed young racial/minority adults face greater risk for depression and anxiety (McKnight-Eily et al., 2021; Mehdi et al., 2020). Our data suggest this is due not only to the infection risks associated with work outside the home during the pandemic, but to victimization by others based on social perceptions that people of color are more likely to be infected with the virus.

Of note, 28% and 22% of participants identified as sexual or gender minorities, respectively. Sexual and gender minority participants reported higher levels of depression and gender minorities also reported higher levels of Coronavirus victimization. As both persons of color and members of sexual and gender minority groups, the current COVID-19 crisis is likely exacerbating mental health stressors among these groups due to healthcare disparities resulting from multiple barriers such as medical mistrust, provider bias, and lack of insurance (Harkness et al., 2020; Ruprecht et al., 2021).

### Coronavirus Victimization Distress, Coronavirus Racial Bias and Mental Health

This study provides evidence of an association between demographic factors, Coronavirus specific victimization and perceived racial bias, and mental health. Participants in our study who were employed, reported prescription insecurity and more COVID-19 health risks reported higher levels of Coronavirus victimization distress and perceived Coronavirus racial bias. These findings underscore the fact, that in addition to health and economic disparities, AIAN, Asian, Black, and Latinx young adults are bearing a disproportionate social burden during the current health crises.

The level of Coronavirus victimization distress and percentage of participants reporting other risk factors did not differ across different racial/ethnic groups, with one exception; Asian and Black respondents reported higher levels of Coronavirus racial bias than AIAN and Latinx. This finding may be explained by political rhetoric and media reports at the time of data collection in April 2020. During the first few months of the pandemic, anti-Asian sentiment was fueled by scapegoating the “China virus.” At the same time, the U.S. was in the early throes of a renewed racial justice movement and had witnessed disproportional COVID-19 infections and morbidity among Black Americans; with outbreaks among Latinx persons and on tribal lands largely ignored by the federal government, mainstream media and online tracking (El Chaar et al., 2020; Natividad, 2020; Owen et al., 2020; Xu et al., 2021). The finding that Asian respondents were in general of higher household income, were less likely to be employed as essential workers or report financial or prescription insecurity demonstrates the pernicious effect of racism on mental health irrespective of other historically documented protective factors (McGee & Thompson, 2015; Paul & Moser, 2009).

Results of the SEM analysis further underscore the impact of racism during the pandemic on mental health. When demographic variables were controlled, the effect of Coronavirus victimization distress on mental health was fully mediated by perceptions of COVID-19 related increases in racial bias across the U.S. These findings may be explained, in part, by prior work on attribution theory, indicating the negative effect on mental health when members of marginalized groups attribute negative social experiences to racial discrimination (Schmitt et al., 2014). However, the CRBS included items on perceptions of Coronavirus related racist social and mass media posts and racial/ethnic discrimination in COVID-19 health care settings that went beyond personal experiences related to bullying and other forms of victimization. Thus, CRBS responses within the context of national statistics documenting increases in racial/ethnic discrimination during the pandemic indicate such attributions were grounded in the reality of COVID-19 related racial strife (Addo, 2020; Dhanani & Franz, 2021; Ruiz et al., 2020).

## Limitations and Future Directions

To our knowledge, this is the first study to examine the relationship to mental health of victimization distress and perceived racial bias directly related to the Coronavirus among AIAN, Asian, Black, and Latinx young adults. However, due to the cross-sectional nature of the survey, we are unable to interpret the results as definitive of causation. It will be important for future studies to examine the longitudinal impact of Coronavirus victimization distress and associated racial bias beliefs on mental health among young adults within these populations. Although the study’s data collection and recruitment methods yielded a geographically diverse national sample of racial/ethnic minority young adults; this methodology does not allow for absolute certainty that inclusion criteria were met and also limited participation to registered respondents with Internet or mobile phone access. Finally, our data also suggest that for some populations, minority stress associated with sexual and gender minority status may have a greater influence on Coronavirus related victimization. It will be fruitful to examine how the intersectionality of race/ethnicity and sexual and gender minority status further exacerbates the effect of Coronavirus related victimization and racial bias on the mental health of young adults of color.

## Conclusions

The mental health of people of color in the United States has been associated with longstanding experiences of racial and ethnic discrimination and systemic bias. This study highlights how the current COVID-19 pandemic has added to these mental health burdens through increases in Coronavirus victimization and perceived racial bias experienced by AIAN, Asian, Black, and Latinx young adults. These findings also demonstrate how an infectious disease crisis can reverse the usual protective effects of employment on mental health, when working people of color are more likely to be employed in settings that increase their exposure to infection and subsequent social bias. In addition, this study underscores the deleterious effects on mental health of pre-existing Coronavirus health risks and associated prescription insecurity and the continuing vulnerabilities associated with financial insecurity and sexual minority status. In addition to ongoing efforts to increase general cultural competencies in mental health services, these findings highlight the urgency of mental health treatments tailored to the specific needs of racial/ethnic minorities during the current and future health crises.

## Data Availability

Once accepted for journal publication, data will be provided on a link established on the primary website https://www.fordham.edu/info/24019/center_for_ethics_education_research/11686/fordham_university_pathways_to_health_study

